# Threshold Effects, Cognitive Decline, and Longitudinal Changes in White Matter Hyperintensity Volume

**DOI:** 10.64898/2025.12.05.25341741

**Authors:** Sarvin Sasannia, Mykola Matsyuk, Shimeng Wang, Jinwei Zhang, Keenan A. Walker, Hyeong-Geol Shin, Richard Leigh, Jerry L. Prince, Lewis C. Becker, Lisa R. Yanek, Nicola J. Armstrong, Peter van Zijl, Dhananjay Vaidya, Linda Knutsson, Paul A. Nyquist

## Abstract

**Background:** Changes in Ischemic White matter hyperintensities volume (WMH) on MRI over time are associated with cognitive decline. We investigated whether changes in WMH volume over time exhibit WMH-normalized volume threshold effects on declining cognitive performance and whether these effects on cognition differ between deep white matter hyperintensities (DWMH) and periventricular white matter hyperintensities (PVWMH).

**Methods:** We followed 339 participants longitudinally from GeneSTAR with brain MRI and neuropsychological testing at baseline and at 13-years (62% female, and 33% Black, mean baseline age 49.7±9.6). WMH were classified as PVWMH (within 2mm of ventricles) or DWMH. Two-segment linear spline regression models using adjusted mixed linear regression identified test-specific thresholds longitudinally beyond which cognitive decline accelerated. Cognitive scores from both timepoints were treated as repeated measures, with WMH included as a time-varying predictor.

**Results:** Declines in motor function and processing speed accelerated beyond thresholds of changing PVWMH and DWMH volumes. For Grooved Pegboard tests, changes in volume were associated with minimal effects below a threshold of changing volume (log-transformed ratio of lesion volume to intracranial volume for: PVWMH −9.42 to −9.29; and DWMH −11.8 to −11.7). Substantial declines in cognitive performance were observed above thresholds of increases in volume (slope differences: PVWMH; 14.5-15.1 seconds per log-unit, p<0.001; and DWMH; 9.54-10.9, p<0.001). Digit Symbol Substitution Test demonstrated paradoxical positive associations below changing volume thresholds (PVWMH; β=6.68, p=0.001 and DWMH; β=6.98, p<0.001), reversing to decline above thresholds of increase in volume for PVWMH (Δβ=−11.2, p<0.001) and DWMH (Δβ=−9.77, p<0.001).

**Conclusion:** Changes in WMH volume exhibit nonlinear threshold effects on changes in cognitive performance over time and differ by anatomic region. Minimal cognitive impact occurred below thresholds, with accelerated declines above. PVWMH demonstrate larger effects on declining cognitive function than DWMH, particularly for motor and processing speed functions and progresses at a faster rate.

## Introduction

Cerebral small vessel disease (cSVD) is a major contributor to cerebrovascular and cognitive morbidity, accounting for about one quarter of ischemic strokes and up to half of dementia cases, and is associated with gait, mood, and neurobehavioral impairments.^1^

A characteristic imaging feature of cSVD is white matter hyperintensities (WMH), which are areas of increased signal on T2-weighted and fluid-attenuated inversion recovery (FLAIR) magnetic resonance imaging (MRI). Their clinical and research relevance has been underscored in the latest cSVD imaging consensus guidelines (STRIVE-2).^2^

WMH are associated with increased risk of cognitive dysfunction.^3^ Their topographic distribution has been demonstrated to influence the cognitive profile of individuals with cSVD.^4,5^ In particular, the distinction between periventricular WMH (PVWMH), located adjacent to the ventricles, and deep WMH (DWMH) has been recognized as an important framework for classifying WMH topography.^6,7^

Growing evidence indicates that the spatial distribution of WMH matters for cognitive outcomes, but its importance may vary depending on the context. PVWMH have been frequently associated with greater cognitive impairment, particularly affecting executive functions, processing speed, and memory.^7,8^ Other studies indicate that, conversely, DWMH contribute to a diffuse reduction in cognitive efficiency,^9^ and some longitudinal data suggest that increasing DWMH load is associated with memory decline.^10^ However, an important unresolved question is whether the relationship between volumetric WMH burden and cognition is linear across all levels of disease severity, or whether threshold effects exist whereby cognitive decline accelerates only after WMH burden exceeds critical levels.

We have systematically examined the clinical characteristics of apparently healthy, middle-aged individuals with cSVD, with a particular focus on PVWMH and DWMH.^11–14^ We first reported a strong association between WMH volume and impaired manual dexterity as measured by the grooved pegboard test (GPBT).^11^ Follow-up analyses linked poor GPBT performance to disrupted white matter microstructural integrity, suggesting tract injury as a mechanistic basis for reduced manual dexterity.^15^ More recently, we extended this line of research in a larger cohort, demonstrating that PVWMH is more strongly associated than DWMH with deficits in manual dexterity and word recall, highlighting distinct cognitive correlates of WMH topography.^16^

Building on these findings, in this study, we sought to characterize the longitudinal trajectories of PVWMH and DWMH over a 13-year period, and to determine whether these trajectories differ by location. Recent study supports the idea that WMH burden may exert threshold-like effects on cognition. Several studies report that once PWMH volume exceeds a certain range, declines in executive function and processing speed become noticeably steeper, whereas the influence of deep WMH is more variable and often less pronounced.^17,18^ These regional differences likely reflect distinct underlying mechanisms; periventricular areas include densely interconnected fiber pathways that may be especially susceptible to injury.^19^ Cognitive performance may appear relatively stable until WMH accumulation reaches a point at which compensatory networks can no longer offset disruption of these tracts. Flexible statistical approaches, including spline and piecewise regression, have been increasingly used to capture these inflection points and have shown that nonlinear patterns better describe the WMH–cognition relationship than traditional linear models,^18,20^ Using linear spline regression models, we investigated whether threshold effects exist in the relationship between WMH accumulation and cognitive decline, and whether these thresholds differ by WMH location and cognitive domain. We hypothesize that both DWMH and PVWMH demonstrate threshold effects on cognition, with domain-specific vulnerabilities whereby PVWMH preferentially affects motor and processing speed domains, while DWMH more strongly influences memory and executive functions. Our hypothesis aligns with earlier cross-sectional observations from our group^16^ and is consistent with longitudinal findings demonstrating that progression of periventricular WMH is more closely tied to deterioration in attention, executive functioning, and processing speed than the progression of deep WMH.^21–23^

## Materials and methods

### Study Population and Design

Participants were recruited from the Genetic Study of Atherosclerosis Risk (GeneSTAR), an ongoing prospective cohort study of asymptomatic first-degree relatives of individuals who developed coronary artery disease before the age of 60 years.^11^ The GeneSTAR study is designed to characterize risk factors for occult cardiovascular and cerebrovascular disease in this population. See descriptions of the study design as previously reported.^11,12,14,15,24^ In this longitudinal analysis, we included 339 participants who underwent both baseline (timepoint 1) and follow-up (timepoint 2) assessments approximately 13 years apart, with brain MRI and cognitive testing at both timepoints. Exclusion criteria included chronic steroid use; life-threatening medical conditions; neurologic disorders that could impair MRI interpretation; presence of implanted metals incompatible with MRI; atrial fibrillation; and symptomatic cardiovascular or cerebrovascular disease. All participants provided written informed consent, and the study was approved by the Johns Hopkins Institutional Review Board (IRB NA_00002856).

### Clinical Assessment

At both timepoints, participants underwent comprehensive clinical evaluation including detailed medical, social, and family history recording and physician examination. Fasting blood samples were collected for cholesterol and glucose measurements. Blood pressure was measured 3-4 times during the screening day. Hypertension was defined as average blood pressure ≥140/90 mmHg, current use of antihypertensive medication, and/or history of physician-diagnosed hypertension. Diabetes was defined as fasting glucose >125 mg/dL, current use of hypoglycemic medication, and/or history of physician-diagnosed diabetes.

### Brain MRI Acquisition

Brain MRI was acquired at both timepoints using 3-Tesla Philips scanners (Philips Healthcare, Best, the Netherlands). Different scanner models were used at each time point.

Baseline, imaging was performed on a 3T Achieva system. The protocol included a T1-weighted magnetization-prepared rapid gradient-echo (MPRAGE) sequence (repetition time [TR] = 10 ms, echo time [TE] = 6 ms, inversion time [TI] = 932 ms, flip angle = 8°, voxel size acquired = 1.00 × 0.99 × 1.00 mm³ (reconstructed: 0.75× 0.75 × 3 mm³), contiguous 1 mm slices, and 2D (multislice) axial turbo spin-echo fluid-attenuated inversion recovery (T2-FLAIR) (TR = 11,000 ms, TE = 68 ms, inversion time [TI] = 2800 ms, voxel size acquired = 0.98 × 0.96 × 3 mm³ (reconstructed: 0.47× 0.47 × 3 mm³), contiguous 3 mm slices.

At follow-up, ~13 years later, imaging was performed on a 3T Ingenia system. The protocol included a 3D MPRAGE sequence, (TR = 6.4 ms, TE = 2.9 ms, TI = 853 ms, flip angle = 9°, voxel size acquired =1.00 × 1.00 × 1.00 mm³ (reconstructed: = 0.98 × 0.98 × 1.0 mm³), compressed sense factor 4, 192 slices, acquisition time = 1 min 59 s) and a 3D T2-FLAIR sequence (TR = 4800 ms, effective TE = 271 ms, TI = 1650 ms, compressed sense factor 4, voxel size acquired = 1.00 × 1.00 × 1.00 mm³ (reconstructed: = 0.98 × 0.98 × 1.0 mm³), 220 slices, acquisition time = 4 min 36 s). Follow-up imaging was performed on different scanner models resulting in protocol differences in sequence implementation (e.g., repetition time and inversion time for T2-FLAIR). These were addressed during preprocessing and segmentation through standardized intensity normalization and harmonization procedures to ensure comparability across timepoints.

### Image Processing and WMH Segmentation

#### Preprocessing Pipeline

MPRAGE, T2-FLAIR, and T2-weighted images underwent standardized preprocessing. First, intensity inhomogeneity correction was performed using N4 bias field correction.^25^ The multi-slice 2D scans were then super-resolved using SMORE (Self-supervised Anti-aliasing and Super-resolution Algorithm).^26,27^ Images were subsequently skull-stripped using automated artificial neural networks and rigidly co-registered to Montreal Neurological Institute (MNI) space.^28^ To reduce domain shift and scanner-related variability across subjects and timepoints, HACA3 intensity and contrast harmonization was applied.^29,30^

#### WMH Segmentation and Classification

White matter hyperintensity lesions were segmented using UNISELF, a domain-adaptable lesion segmentation model.^30,31^ The segmentation pipeline generated lesion masks using a U-Net architecture, which were then divided into connected components using 18-connectivity.^30^ Brain segmentation was performed using SLANT (Spatially Localized Atlas Network Tiles) to generate whole-brain anatomical masks.^32^ Lesions were classified based on their proximity to the ventricles: components intersecting with the region within 2mm of the ventricles were classified as PVWMH, while remaining lesions were classified as DWMH.

#### Longitudinal Registration and Temporal Correspondence

To account for anatomical changes between timepoints and ensure accurate longitudinal comparison, we implemented a comprehensive registration strategy. The T1 image from baseline was registered to MNI space template using standard linear registration^28^. T2-FLAIR and T2 images from baseline were then registered to the aligned T1 image. For follow-up, the T1 image was registered to the baseline T1 image, followed by registration of the follow-up T2-FLAIR to the follow-up T1.

Deformable registration between timepoints was performed using Voxel Morph to capture subtle anatomical changes over the 13-year interval.^33^ The lesion-filled T1 image from baseline was registered to the T1 image at follow-up, and the corresponding lesion mask was warped accordingly using the computed transformation fields.^34^ Lesions across timepoints were cross-referenced to establish temporal correspondences and track individual lesion evolution.^35^ The lesion classification mask at follow-up was refined using lesion information from baseline to maintain consistency in PVWMH/DWMH classification. Lesion progression and regression were computed by comparing the warped lesion mask from baseline to that of follow-up, enabling precise quantification of volumetric changes. Examples of WMH segmentation and longitudinal tracking are shown in Figure 1.

**Figure 1.**
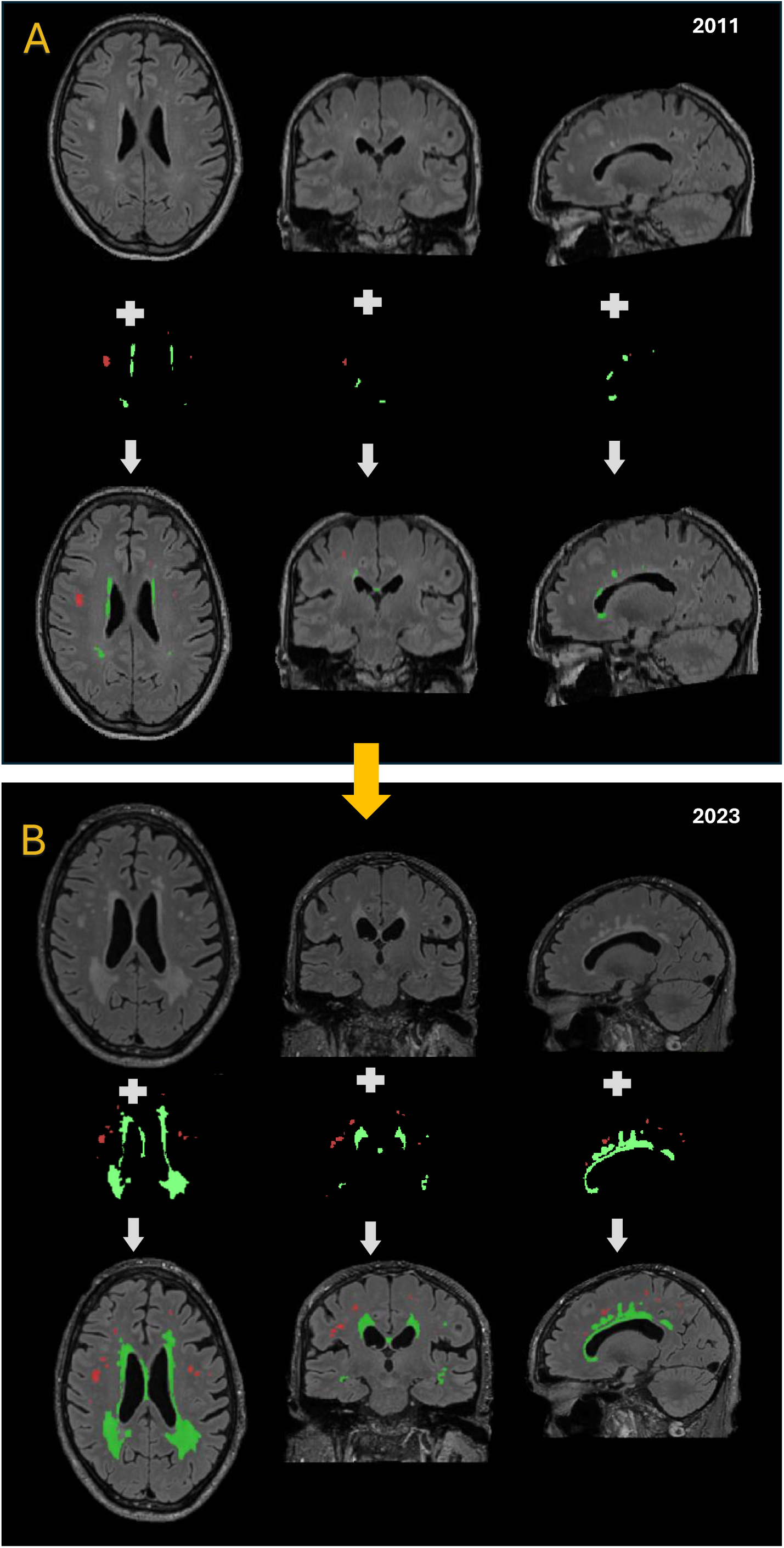
Longitudinal progression of WMH. FLAIR MRI scans from one participant are shown at two timepoints: **(A)** in 2011 and **(B)** in 2023. Each row displays axial, coronal, and sagittal views to demonstrate the spatial distribution of WMH. Green masks indicate periventricular white matter hyperintensities (PVWMH), while red masks indicate deep white matter hyperintensities (DWMH). The 2011 scan was acquired at Johns Hopkins Hospital, and the 2023 scan at the Kennedy Krieger Institute

### Cognitive Assessment

A comprehensive neuropsychological battery was administered by trained personnel on the same day as the brain MRI. The cognitive assessment battery was expanded between timepoints to include additional measures of cognitive function. Cognitive tests administered at both timepoints included: Mini-Mental State Examination (MMSE) for global cognitive screening assessing orientation, memory, attention, language, and praxis (total score 0-30); Digit Span Test for verbal short-term and working memory assessment with forward and backward conditions, plus total score, measuring longest accurately repeated digit sequence; Delayed Word Recall Test for episodic memory assessment with multiple components including short delay, long delay, and immediate recall (Delayed Word Recall 1), scored by number of correctly recalled words; Digit Symbol Substitution Test for assessment of processing speed, attention, and visuomotor function with both total correct and total incorrect components; Phonemic Word Fluency (F/A/S) test of word retrieval and executive control, requiring participants to generate words beginning with specific letters within 1 minute; and Grooved Pegboard Test (GPBT), a measure of manual dexterity requiring placement of 25 pegs as quickly as possible, performed separately for dominant and non-dominant hands, with both completion time and performance scores recorded.

Additional cognitive tests administered only at follow-up included: Montreal Cognitive Assessment (MoCA) for enhanced global cognitive screening with greater sensitivity to mild cognitive impairment; Craft Story Tests for narrative memory assessment with immediate and delayed recall components for both verbatim and paraphrase conditions; Benson Complex Figure Test for visuospatial construction and visual memory assessment; Category Fluency Tests for semantic fluency assessment using animal and vegetable categories; Trail Making Tests A and B for visual attention, processing speed, and executive function assessment; Benson Delayed Total for delayed recall component of the Benson Complex Figure Test; and Multilingual Naming Test (MINT) for confrontation naming assessment across multiple languages.

### Statistical Analysis

All WMH volumes were normalized by dividing intracranial volume measured at follow-up, then log-transformed using the formula log (x + 0.5 × minimum positive value) to reduce right skewness and facilitate comparison across WMH subtypes. The primary analysis employed two-segment linear spline regression models to characterize threshold effects in WMH-cognition relationships. To identify potential threshold effects where the relationship between WMH burden and cognitive performance changes at specific WMH levels, we employed two-segment linear spline mixed linear regression models to determine longitudinal changes. Threshold values (knots) for each cognitive test and WMH type combination were determined by first fitting exploratory quadratic regression models to baseline cross-sectional data. The inflection point from each quadratic model was calculated as: knot = −β_linear/(2×β_quadratic), presenting the WMH level at which the slope of the WMH–cognition relationship is expected to change.

Quadratic model results supported the presence of potential threshold effects. As an additional validation step, we conducted a grid search across WMH values and compared competing models using AIC and BIC criteria. This analysis identified thresholds that followed the same pattern as those estimated from the quadratic models.

These data-driven knot values were then incorporated into the final two-segment linear spline models with the following specification:

### Cognitive Score ~ β_1_(WMH if WMH ≤ knot) + β_2_(WMH - knot if WMH > knot) + covariates

Where β_1_ represents the slope below the threshold and β_2_ represents the slope above the threshold. The difference between slopes (Δβ = β_2_ - β_1_) was tested to determine whether the WMH-cognition relationship significantly differs across the threshold. Separate models were constructed for PVWMH, DWMH, and total WMH, with each model adjusted for age at baseline, sex, race, education, systolic blood pressure, and family clustering using robust standard errors. A significant Δβ indicates the presence of a threshold effect, with cognitive decline accelerating (or decelerating) above the identified WMH burden level.

In these analyses, cognitive test scores at baseline and 13-year follow-up served as repeated outcome measures. We did not compute explicit change scores; instead, the mixed-effects model used cognitive scores at each timepoint, with WMH (also measured at both timepoints) included as a time-varying predictor. Thus, ‘change’ or ‘decline’ refers to differences in cognitive performance captured across the two repeated assessments.

Cross-sectional associations between WMH volume (total, PVWMH, and DWMH) were measured at follow-up only and concurrent cognitive performance at the second timepoint using linear regression models adjusted for the same covariates with single point estimate of MOCA score (see Supplementary Table 1).

Given that 26 cognitive tests were evaluated and many were correlated within domains, we controlled for multiple comparisons using false discovery rate (FDR) correction applied separately within each cognitive domain. All analyses were performed using R statistical software (version 4.5.1 or later). Spline regression models were implemented using the lspline package.

### Data Availability

The data supporting the findings of this study are available from the GeneSTAR steering committee upon reasonable request, subject to appropriate data sharing agreements to protect participant privacy.

## Results

### Participant Characteristics

A total of 350 participants from the GeneSTAR cohort completed baseline assessments (mean age 49.7±9.6 years, 62% female) and follow-up evaluations approximately 13 years later (mean age 63.2±9.7 years). Complete baseline WMH data were available for 339 participants, with 11 participants excluded due to missing or inadequate T2-FLAIR or T2-weighted imaging sequences. The cohort was predominantly European American (66%) and Black (33%), with a mean education level of 14.5±2.6 years. Baseline WMH volume was modest, with median total WMH of 686 mm³ (IQR: 364–1207), median PVWMH of 545 mm³ (IQR: 258–959), and median DWMH of 107 mm³ (IQR: 17–281). Detailed baseline and follow-up demographic and clinical characteristics are presented in Table 1, and cognitive test performance at both timepoints is summarized in Table 2.

**Table 1.**
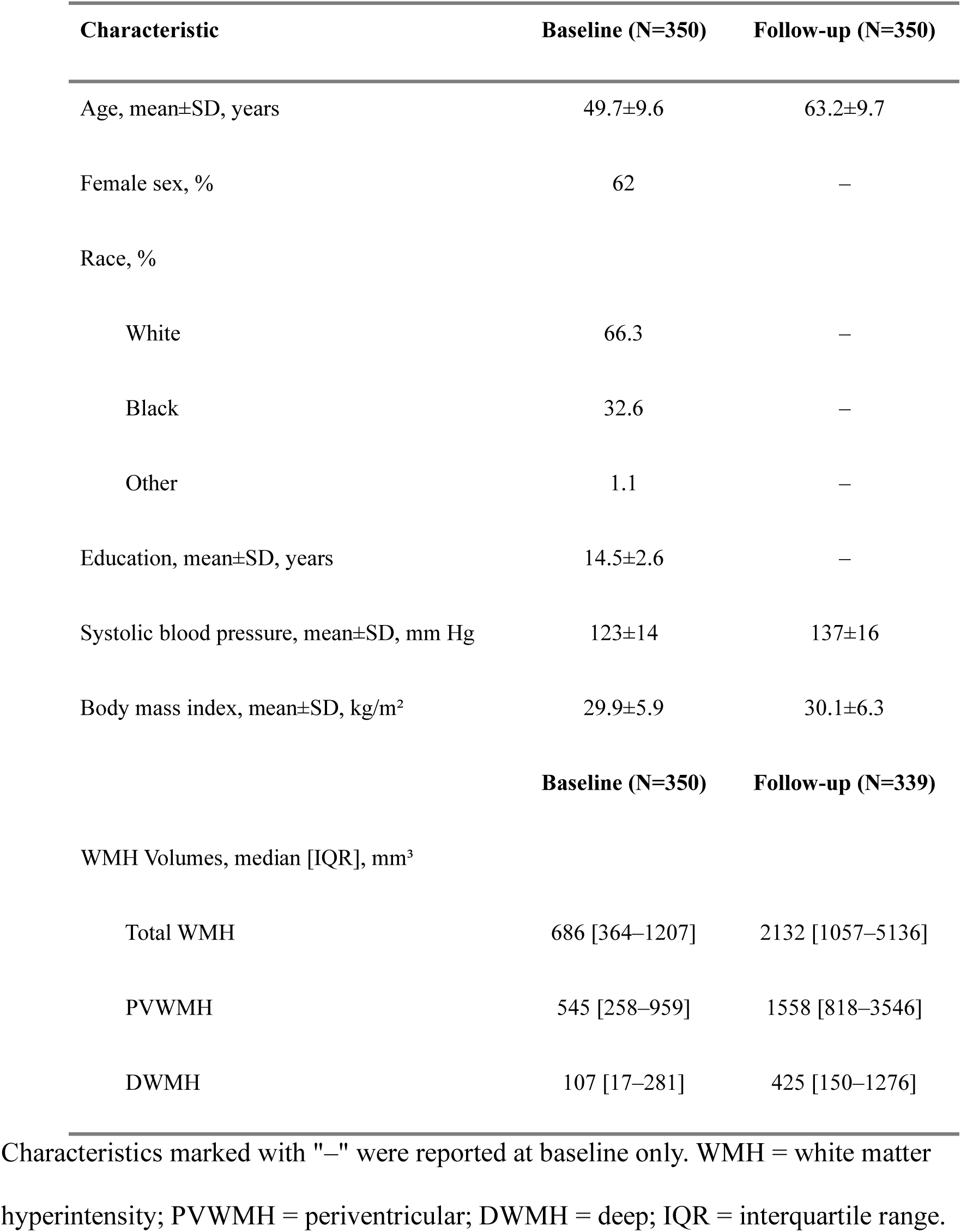
Baseline and Follow-up Characteristics.

| Characteristic | Baseline (N=350) | Follow-up (N=350) |
| --- | --- | --- |
| Age, mean±SD, years | 49.7±9.6 | 63.2±9.7 |
| Female sex, % | 62 | — |
| Race, % |  |  |
| White | 66.3 | — |
| Black | 32.6 | — |
| Other | 1.1 | — |
| Education, mean±SD, years | 14.5±2.6 | — |
| Systolic blood pressure, mean±SD, mm Hg | 123±14 | 137±16 |
| Body mass index, mean±SD, kg/m<sup>2</sup> | 29.9±5.9 | 30.1±6.3 |
|  | **Baseline (N=350)** | **Follow-up (N=339)** |
| WMH Volumes, median [IQR], mm<sup>3</sup> |  |  |
| Total WMH | 686 [364–1207] | 2132 [1057–5136] |
| PVWMH | 545 [258–959] | 1558 [818–3546] |
| DWMH | 107 [17–281] | 425 [150–1276] |

Characteristics marked with "—" were reported at baseline only. WMH = white matter hyperintensity; PVWMH = periventricular; DWMH = deep; IQR = interquartile range.

**Table 2.** Cognitive Test Performance at Baseline and Follow-up.

| Cognitive Test | Baseline Score | Follow-up Score |
| --- | --- | --- |
| MMSE | 29 [28–30]; N=348 | 30 [29–30]; N=348 |
| MoCA | – | 27 [25–29]; N=336 |
| Delayed word recall – short form | 7 [6–8]; N=349 | 6 [5–8]; N=347 |
| Delayed word recall – long form | 7 [6–8]; N=346 | 6 [5–8]; N=345 |
| Delayed word recall – immediate | 10 [10–10]; N=349 | 10 [9–10]; N=349 |
| Craft Story – immediate verbatim | – | 21.2±8; N=350 |
| Craft Story – immediate paraphrase | – | 14.19±4.12; N=350 |
| Craft Story – delayed verbatim | – | 18.83±8.34; N=349 |
| Craft Story – delayed paraphrase | – | 13.18±4.53; N=349 |
| Digit symbol substitution – correct | 55.2±11.5; N=348 | 45.2±12.7; N=350 |
| Digit symbol substitution – incorrect | 0.11±0.42; N=348 | 0.20±0.62; N=321 |
| Trail Making Test A | – | 32.9±19.9; N=350 |
| Trail Making Test B | – | 90.4±79.3; N=348 |
| Word fluency | 41.0±11.9; N=346 | 40.7±12.7; N=349 |
| Category fluency – animals | – | 20.3±5.4; N=350 |
| Category fluency – vegetables | – | 14.1±3.9; N=350 |
| MINT | – | 29.7±2.2; N=349 |
| Benson figure – copy | – | 15.5±1.5; N=349 |
| Benson figure – delayed | – | 11.6±3.5; N=338 |

| **Cognitive Test** | **Baseline Score** | **Follow-up Score** |
| --- | --- | --- |
| Digit span – forward | 10.5±2.3; N=349 | 12.3±2.3; N=350 |
| Digit span – backward | 6.63±2.67; N=349 | 7.31±2.69; N=350 |
| Digit span – total | 17.2±4.4; N=349 | 19.6±4.5; N=348 |
| Grooved pegboard – dominant hand time | 76.4±15.2; N=345 | 92.1±52.07; N=341 |
| Grooved pegboard – dominant hand score | 101.7±15.3; N=344 | 117.6±52.8; N=335 |
| Grooved pegboard – nondominant hand time | 84.4±19.8; N=345 | 100.1±64.4; N=344 |
| Grooved pegboard – nondominant hand score | 109.9±20.0; N=344 | 125.5±64.8; N=343 |

Cognitive Test Performance at Baseline and Follow-up Cognitive. performance at baseline and 13-year follow-up. Values shown as median [IQR] or mean±SD. N = number of participants completing each test. Tests with "-" were only administered at follow-up. MMSE = Mini-Mental State Examination; MoCA = Montreal Cognitive Assessment; MINT = Multilingual Naming Test.

### Normalized Lesion Volume Threshold Effects in WMH-Cognition Relationships

We modeled cognition across two repeated timepoints, allowing us to estimate how WMH burden relates to cognitive performance below and above thresholds. The term ‘decline’ reflects differences observed across the repeated measures rather than a separate change variable.

Two-segment linear spline regression models revealed significant threshold effects in the relationships between normalized WMH volume and cognitive performance across multiple domains. The presence and magnitude of these threshold effects varied by cognitive domain and WMH location (Table 3, Figure 2, Supplemental Figure S1). The most robust and consistent threshold effects were observed for tests of manipulative manual dexterity. All four grooved pegboard measures (time and score for both dominant and nondominant hands) demonstrated highly significant slope differences (Δβ, p<0.001) across the threshold for both PVWMH and total WMH, indicating that the detrimental effects of WMH on motor function accelerate markedly after exceeding specific burden levels.

**Figure 2.**
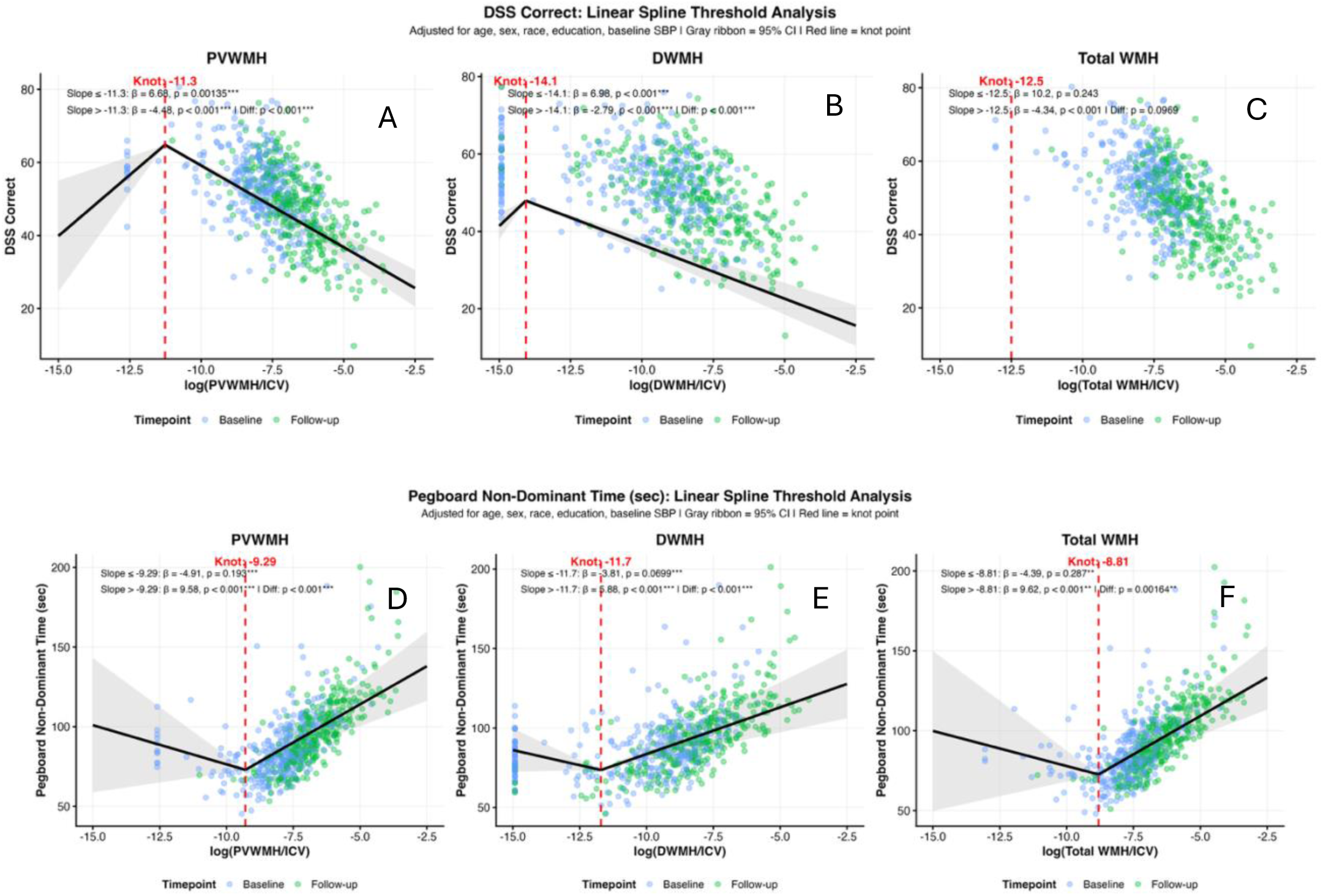
Two-segment linear spline fits demonstrating threshold effects of white matter hyperintensities on cognitive performance. Panels A-C show processing speed (Digit Symbol Substitution Test correct responses) with paradoxical positive associations below threshold reversing to substantial decline above threshold for periventricular (PVWMH), deep (DWMH), and total white matter hyperintensities. Panels D-F show motor function (Grooved Pegboard non-dominant hand time) with minimal effects below threshold and accelerated decline above threshold across all WMH types. Vertical lines indicate threshold points where cognitive decline accelerates. PVWMH = periventricular white matter hyperintensities; DWMH = deep white matter hyperintensities; WMH = white matter hyperintensities.

**Table 3.**
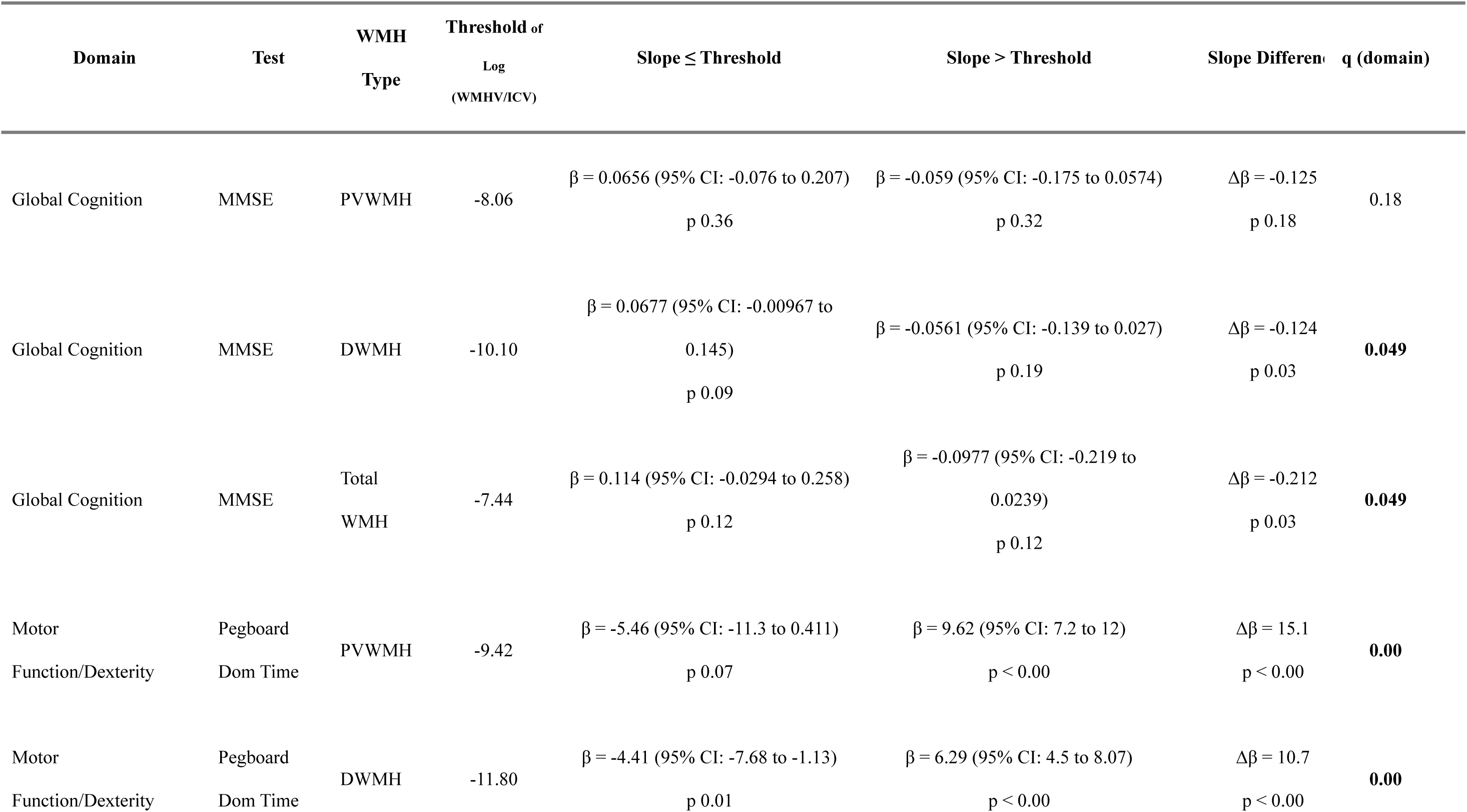
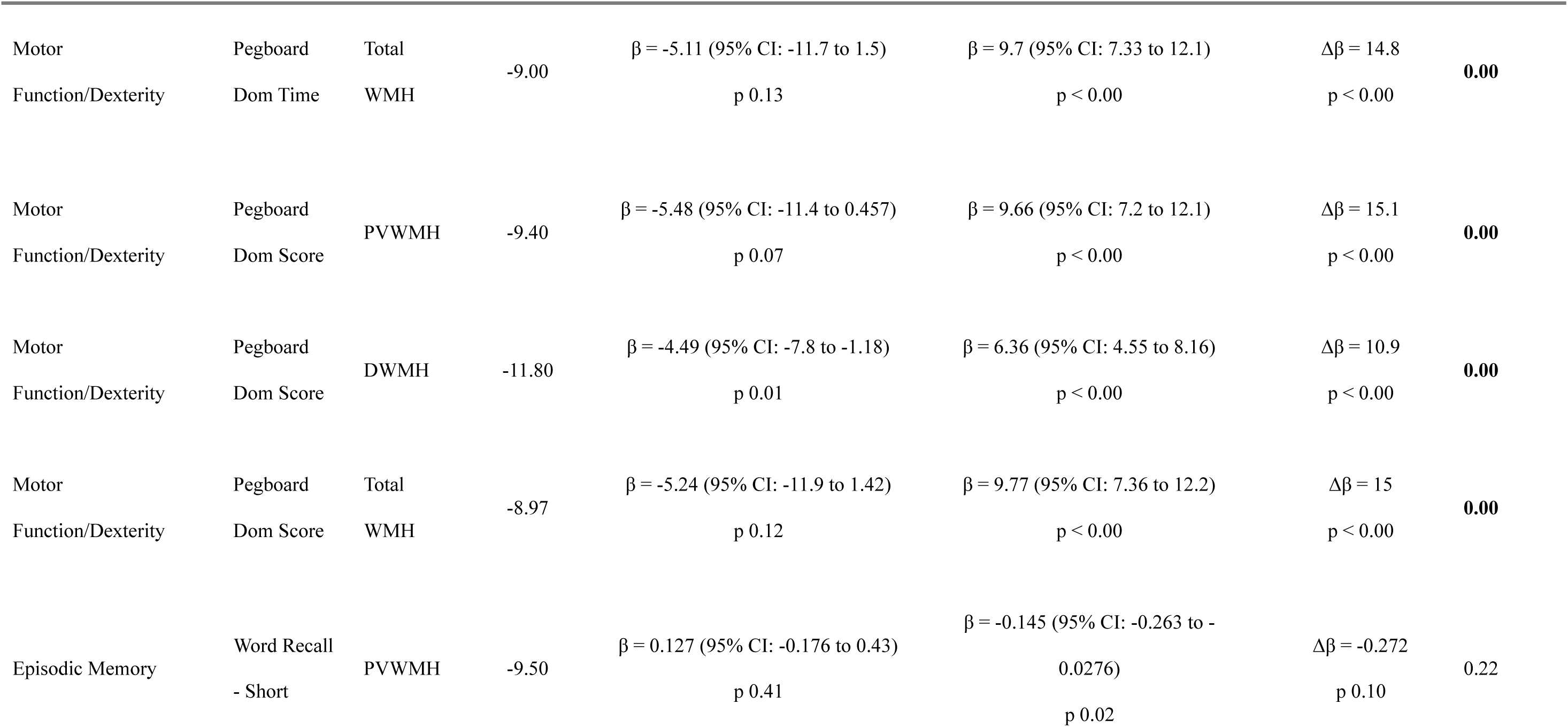
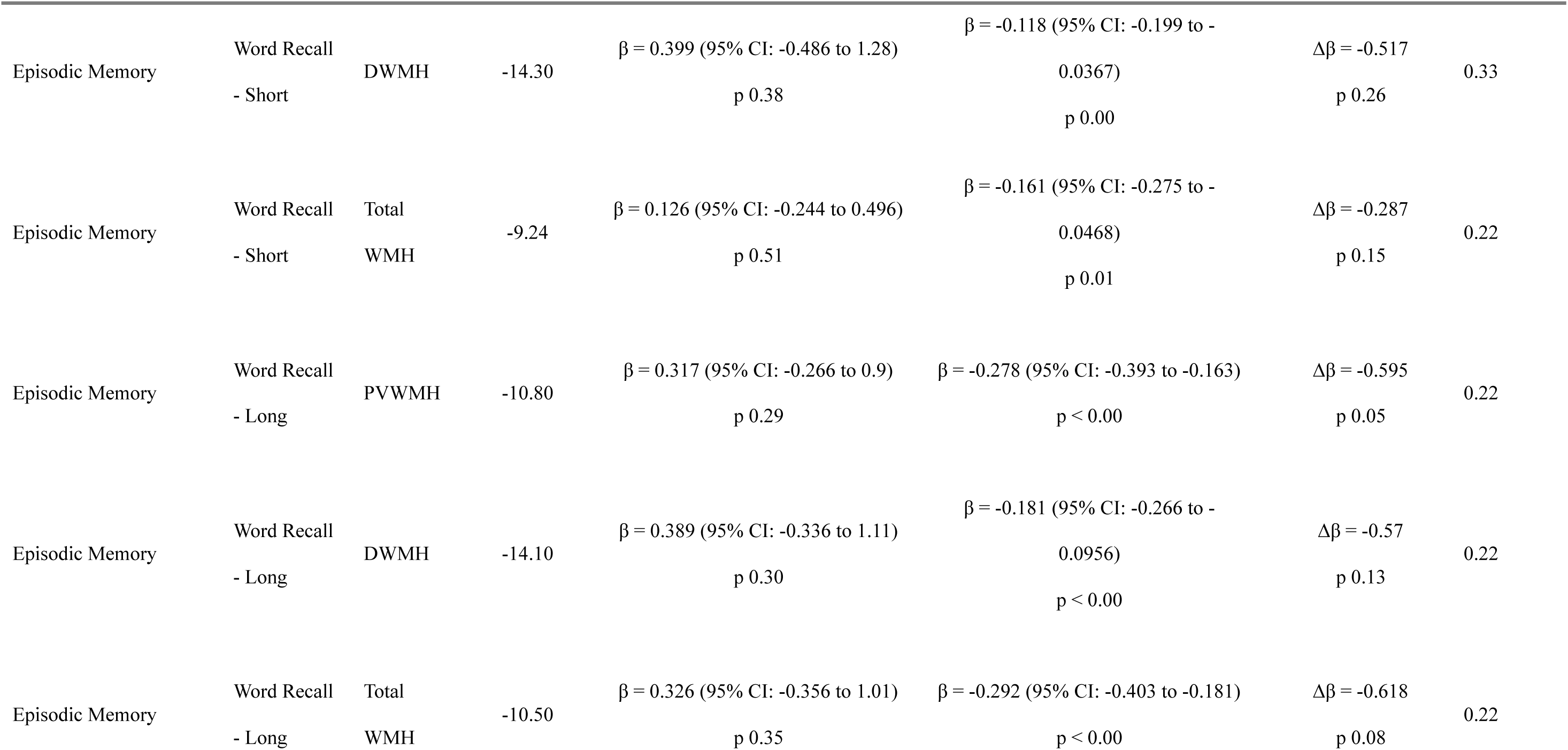
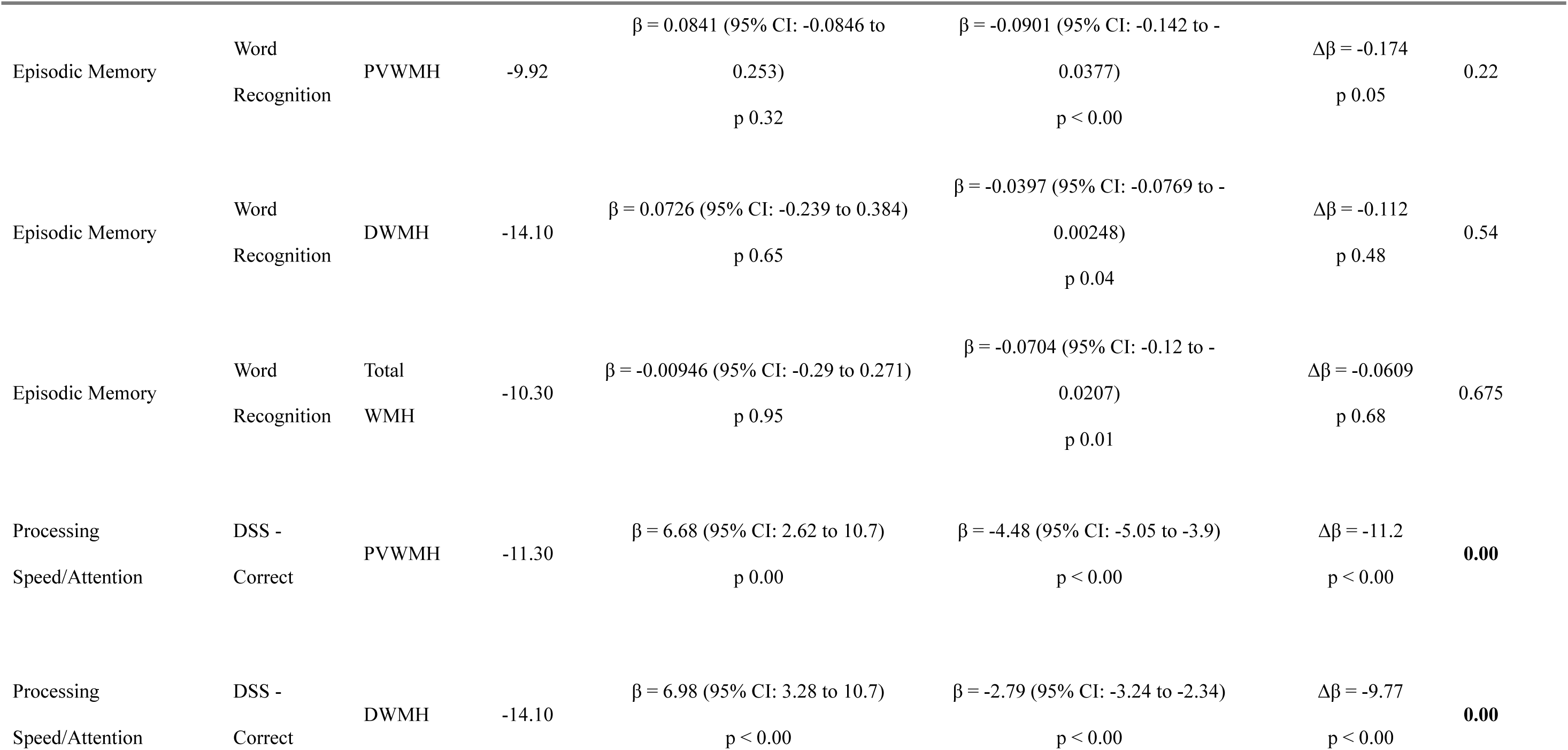
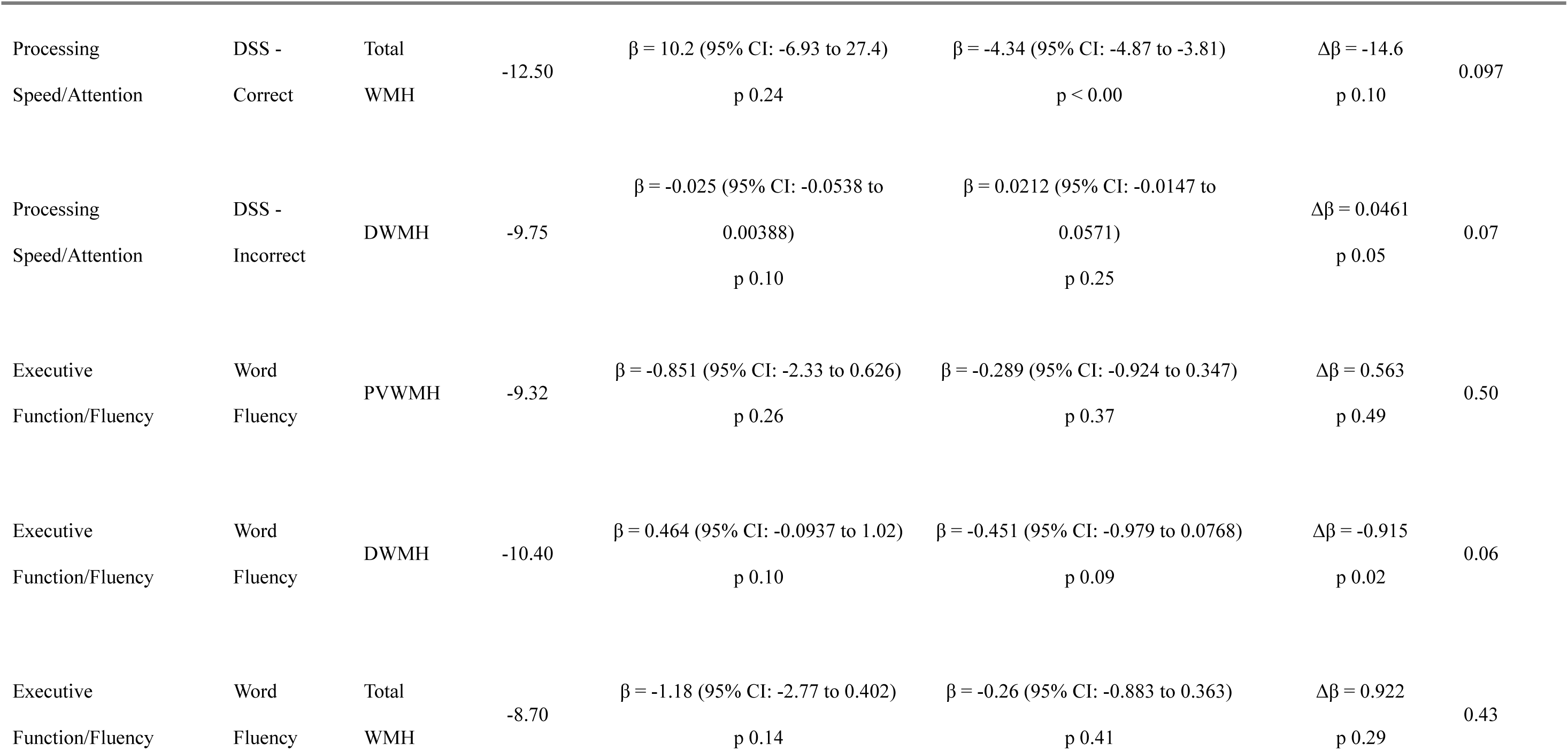
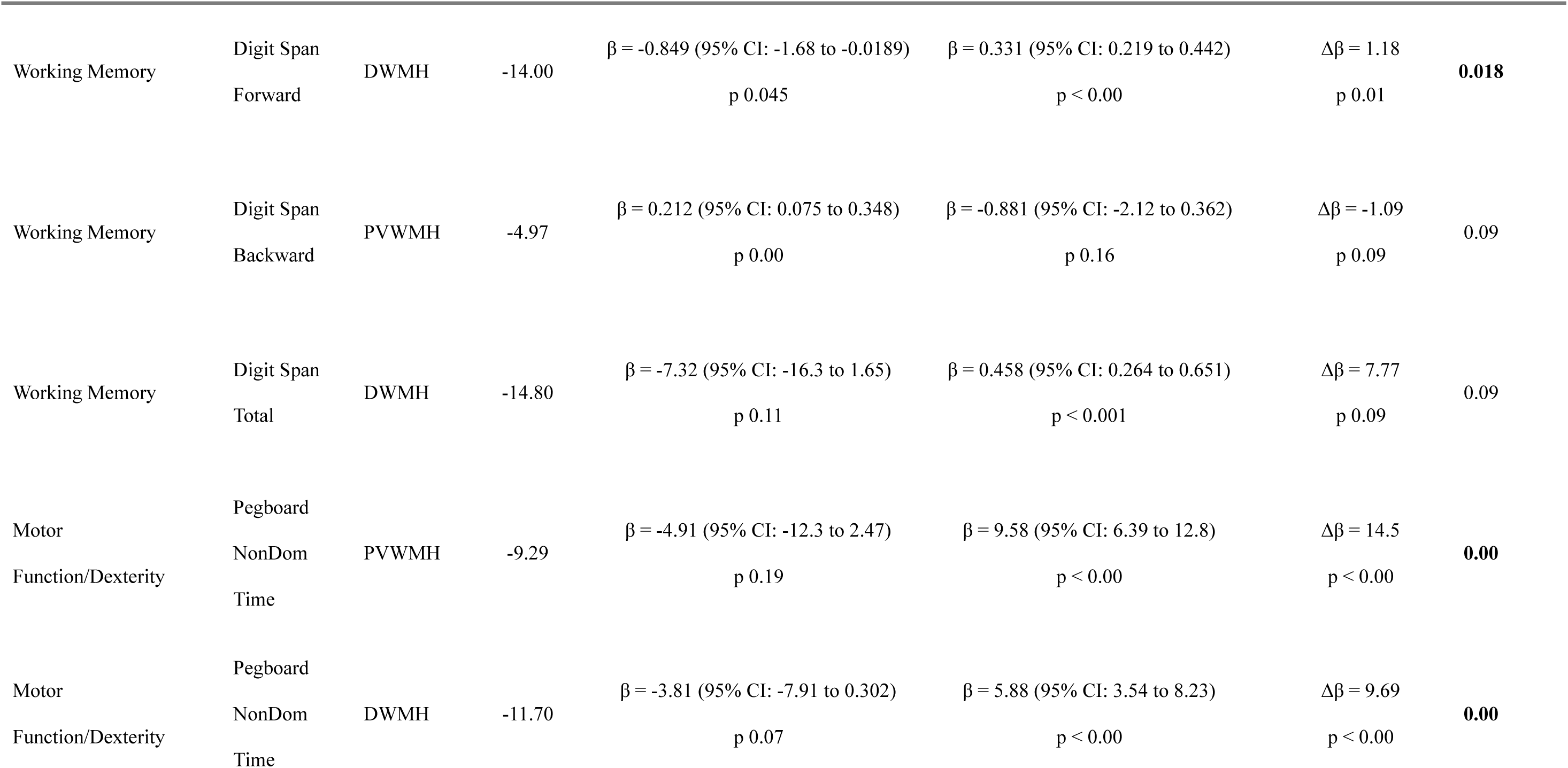
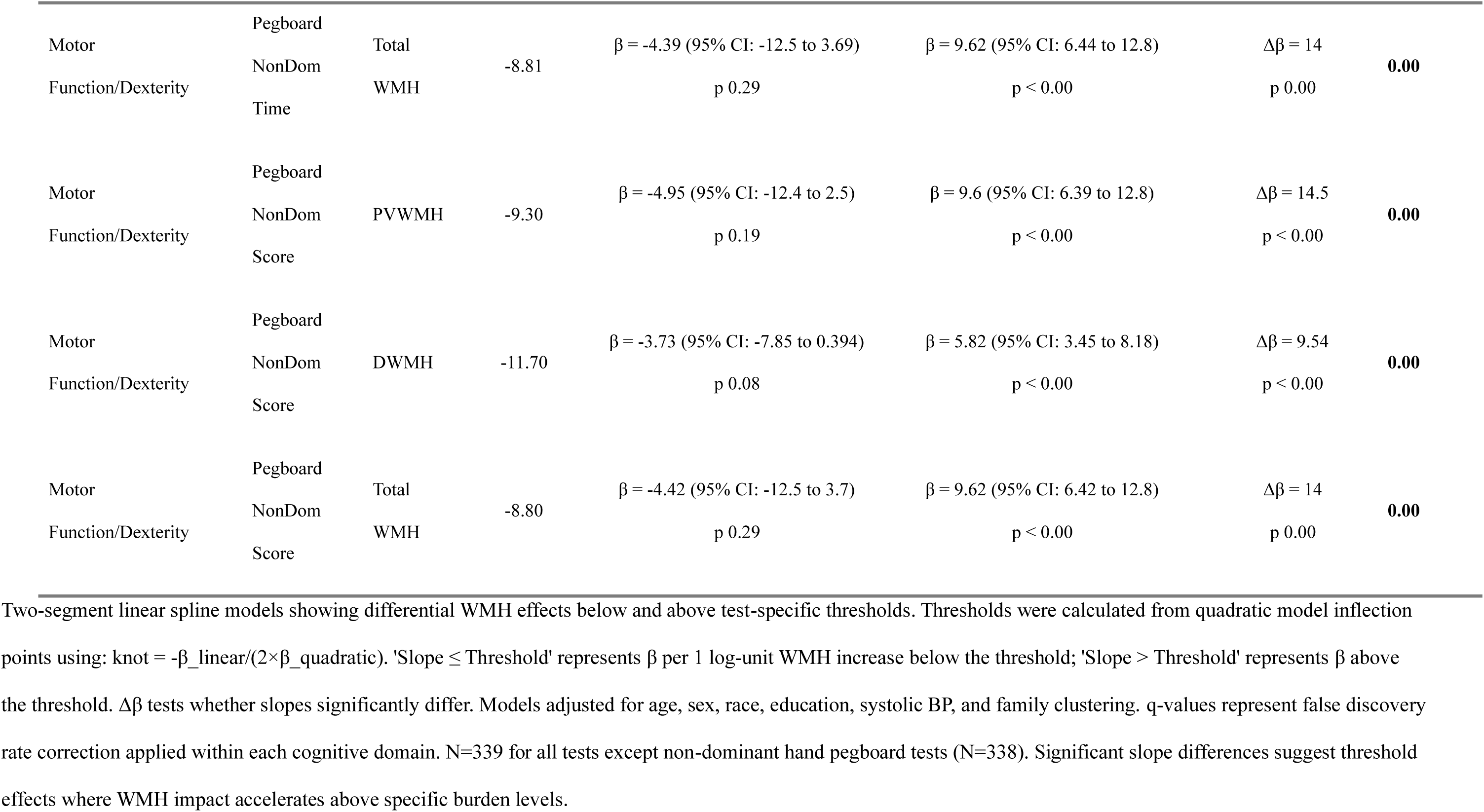
Linear Spline Analysis of WMH Effects on Cognitive Performance.

| Domain | Test | WMH Type | Threshold of Log (WMHV/ICV) | Slope ≤ Threshold | Slope $>$ Threshold | Slope Difference | q (domain) |
| --- | --- | --- | --- | --- | --- | --- | --- |
| Global Cognition | MMSE | PVWMH | -8.06 | $\beta = 0.0656$ (95% CI: -0.076 to 0.207)<br>p 0.36 | $\beta = -0.059$ (95% CI: -0.175 to 0.0574)<br>p 0.32 | $\Delta\beta = -0.125$<br>p 0.18 | 0.18 |
| Global Cognition | MMSE | DWMH | -10.10 | $\beta = 0.0677$ (95% CI: -0.00967 to 0.145)<br>p 0.09 | $\beta = -0.0561$ (95% CI: -0.139 to 0.027)<br>p 0.19 | $\Delta\beta = -0.124$<br>p 0.03 | **0.049** |
| Global Cognition | MMSE | Total WMH | -7.44 | $\beta = 0.114$ (95% CI: -0.0294 to 0.258)<br>p 0.12 | $\beta = -0.0977$ (95% CI: -0.219 to 0.0239)<br>p 0.12 | $\Delta\beta = -0.212$<br>p 0.03 | **0.049** |
| Motor Function/Dexterity | Pegboard Dom Time | PVWMH | -9.42 | $\beta = -5.46$ (95% CI: -11.3 to 0.411)<br>p 0.07 | $\beta = 9.62$ (95% CI: 7.2 to 12)<br>p $< 0.00$ | $\Delta\beta = 15.1$<br>p $< 0.00$ | **0.00** |
| Motor Function/Dexterity | Pegboard Dom Time | DWMH | -11.80 | $\beta = -4.41$ (95% CI: -7.68 to -1.13)<br>p 0.01 | $\beta = 6.29$ (95% CI: 4.5 to 8.07)<br>p $< 0.00$ | $\Delta\beta = 10.7$<br>p $< 0.00$ | **0.00** |

| Domain | Test | WMH Type | Threshold of Log (WMHV/ICV) | Slope ≤ Threshold | Slope > Threshold | Slope Difference | q (domain) |
| --- | --- | --- | --- | --- | --- | --- | --- |
| Motor Function/Dexterity | Pegboard Dom Time | Total WMH | -9.00 | $\beta = -5.11$ (95% CI: -11.7 to 1.5)<br>p 0.13 | $\beta = 9.7$ (95% CI: 7.33 to 12.1)<br>p < 0.00 | $\Delta\beta = 14.8$<br>p < 0.00 | **0.00** |
| Motor Function/Dexterity | Pegboard Dom Score | PVWMH | -9.40 | $\beta = -5.48$ (95% CI: -11.4 to 0.457)<br>p 0.07 | $\beta = 9.66$ (95% CI: 7.2 to 12.1)<br>p < 0.00 | $\Delta\beta = 15.1$<br>p < 0.00 | **0.00** |
| Motor Function/Dexterity | Pegboard Dom Score | DWMH | -11.80 | $\beta = -4.49$ (95% CI: -7.8 to -1.18)<br>p 0.01 | $\beta = 6.36$ (95% CI: 4.55 to 8.16)<br>p < 0.00 | $\Delta\beta = 10.9$<br>p < 0.00 | **0.00** |
| Motor Function/Dexterity | Pegboard Dom Score | Total WMH | -8.97 | $\beta = -5.24$ (95% CI: -11.9 to 1.42)<br>p 0.12 | $\beta = 9.77$ (95% CI: 7.36 to 12.2)<br>p < 0.00 | $\Delta\beta = 15$<br>p < 0.00 | **0.00** |
| Episodic Memory | Word Recall - Short | PVWMH | -9.50 | $\beta = 0.127$ (95% CI: -0.176 to 0.43)<br>p 0.41 | $\beta = -0.145$ (95% CI: -0.263 to -0.0276)<br>p 0.02 | $\Delta\beta = -0.272$<br>p 0.10 | 0.22 |
| Episodic Memory | Word Recall - Short | DWMH | -14.30 | $\beta = 0.399$ (95% CI: -0.486 to 1.28)<br>p 0.38 | $\beta = -0.118$ (95% CI: -0.199 to -0.0367)<br>p 0.00 | $\Delta\beta = -0.517$<br>p 0.26 | 0.33 |
| Episodic Memory | Word Recall - Short | Total WMH | -9.24 | $\beta = 0.126$ (95% CI: -0.244 to 0.496)<br>p 0.51 | $\beta = -0.161$ (95% CI: -0.275 to -0.0468)<br>p 0.01 | $\Delta\beta = -0.287$<br>p 0.15 | 0.22 |
| Episodic Memory | Word Recall - Long | PVWMH | -10.80 | $\beta = 0.317$ (95% CI: -0.266 to 0.9)<br>p 0.29 | $\beta = -0.278$ (95% CI: -0.393 to -0.163)<br>p < 0.00 | $\Delta\beta = -0.595$<br>p 0.05 | 0.22 |
| Episodic Memory | Word Recall - Long | DWMH | -14.10 | $\beta = 0.389$ (95% CI: -0.336 to 1.11)<br>p 0.30 | $\beta = -0.181$ (95% CI: -0.266 to -0.0956)<br>p < 0.00 | $\Delta\beta = -0.57$<br>p 0.13 | 0.22 |
| Episodic Memory | Word Recall - Long | Total WMH | -10.50 | $\beta = 0.326$ (95% CI: -0.356 to 1.01)<br>p 0.35 | $\beta = -0.292$ (95% CI: -0.403 to -0.181)<br>p < 0.00 | $\Delta\beta = -0.618$<br>p 0.08 | 0.22 |
| Episodic Memory | Word Recognition | PVWMH | -9.92 | $\beta = 0.0841$ (95% CI: -0.0846 to 0.253)<br>p 0.32 | $\beta = -0.0901$ (95% CI: -0.142 to -0.0377)<br>p < 0.00 | $\Delta\beta = -0.174$<br>p 0.05 | 0.22 |
| Episodic Memory | Word Recognition | DWMH | -14.10 | $\beta = 0.0726$ (95% CI: -0.239 to 0.384)<br>p 0.65 | $\beta = -0.0397$ (95% CI: -0.0769 to -0.00248)<br>p 0.04 | $\Delta\beta = -0.112$<br>p 0.48 | 0.54 |
| Episodic Memory | Word Recognition | Total WMH | -10.30 | $\beta = -0.00946$ (95% CI: -0.29 to 0.271)<br>p 0.95 | $\beta = -0.0704$ (95% CI: -0.12 to -0.0207)<br>p 0.01 | $\Delta\beta = -0.0609$<br>p 0.68 | 0.675 |
| Processing Speed/Attention | DSS - Correct | PVWMH | -11.30 | $\beta = 6.68$ (95% CI: 2.62 to 10.7)<br>p 0.00 | $\beta = -4.48$ (95% CI: -5.05 to -3.9)<br>p < 0.00 | $\Delta\beta = -11.2$<br>p < 0.00 | **0.00** |
| Processing Speed/Attention | DSS - Correct | DWMH | -14.10 | $\beta = 6.98$ (95% CI: 3.28 to 10.7)<br>p < 0.00 | $\beta = -2.79$ (95% CI: -3.24 to -2.34)<br>p < 0.00 | $\Delta\beta = -9.77$<br>p < 0.00 | **0.00** |
| Processing Speed/Attention | DSS - Correct | Total WMH | -12.50 | $\beta = 10.2$ (95% CI: -6.93 to 27.4)<br>p 0.24 | $\beta = -4.34$ (95% CI: -4.87 to -3.81)<br>p < 0.00 | $\Delta\beta = -14.6$<br>p 0.10 | 0.097 |
| Processing Speed/Attention | DSS - Incorrect | DWMH | -9.75 | $\beta = -0.025$ (95% CI: -0.0538 to 0.00388)<br>p 0.10 | $\beta = 0.0212$ (95% CI: -0.0147 to 0.0571)<br>p 0.25 | $\Delta\beta = 0.0461$<br>p 0.05 | 0.07 |
| Executive Function/Fluency | Word Fluency | PVWMH | -9.32 | $\beta = -0.851$ (95% CI: -2.33 to 0.626)<br>p 0.26 | $\beta = -0.289$ (95% CI: -0.924 to 0.347)<br>p 0.37 | $\Delta\beta = 0.563$<br>p 0.49 | 0.50 |
| Executive Function/Fluency | Word Fluency | DWMH | -10.40 | $\beta = 0.464$ (95% CI: -0.0937 to 1.02)<br>p 0.10 | $\beta = -0.451$ (95% CI: -0.979 to 0.0768)<br>p 0.09 | $\Delta\beta = -0.915$<br>p 0.02 | 0.06 |
| Executive Function/Fluency | Word Fluency | Total WMH | -8.70 | $\beta = -1.18$ (95% CI: -2.77 to 0.402)<br>p 0.14 | $\beta = -0.26$ (95% CI: -0.883 to 0.363)<br>p 0.41 | $\Delta\beta = 0.922$<br>p 0.29 | 0.43 |
| Working Memory | Digit Span Forward | DWMH | -14.00 | $\beta = -0.849$ (95% CI: -1.68 to -0.0189)<br>p 0.045 | $\beta = 0.331$ (95% CI: 0.219 to 0.442)<br>p < 0.00 | $\Delta\beta = 1.18$<br>p 0.01 | **0.018** |
| Working Memory | Digit Span Backward | PVWMH | -4.97 | $\beta = 0.212$ (95% CI: 0.075 to 0.348)<br>p 0.00 | $\beta = -0.881$ (95% CI: -2.12 to 0.362)<br>p 0.16 | $\Delta\beta = -1.09$<br>p 0.09 | 0.09 |
| Working Memory | Digit Span Total | DWMH | -14.80 | $\beta = -7.32$ (95% CI: -16.3 to 1.65)<br>p 0.11 | $\beta = 0.458$ (95% CI: 0.264 to 0.651)<br>p < 0.001 | $\Delta\beta = 7.77$<br>p 0.09 | 0.09 |
| Motor Function/Dexterity | Pegboard NonDom Time | PVWMH | -9.29 | $\beta = -4.91$ (95% CI: -12.3 to 2.47)<br>p 0.19 | $\beta = 9.58$ (95% CI: 6.39 to 12.8)<br>p < 0.00 | $\Delta\beta = 14.5$<br>p < 0.00 | **0.00** |
| Motor Function/Dexterity | Pegboard NonDom Time | DWMH | -11.70 | $\beta = -3.81$ (95% CI: -7.91 to 0.302)<br>p 0.07 | $\beta = 5.88$ (95% CI: 3.54 to 8.23)<br>p < 0.00 | $\Delta\beta = 9.69$<br>p < 0.00 | **0.00** |
| Motor Function/Dexterity | Pegboard NonDom Time | Total WMH | -8.81 | $\beta = -4.39$ (95% CI: -12.5 to 3.69)<br>p 0.29 | $\beta = 9.62$ (95% CI: 6.44 to 12.8)<br>p < 0.00 | $\Delta\beta = 14$<br>p 0.00 | **0.00** |
| Motor Function/Dexterity | Pegboard NonDom Score | PVWMH | -9.30 | $\beta = -4.95$ (95% CI: -12.4 to 2.5)<br>p 0.19 | $\beta = 9.6$ (95% CI: 6.39 to 12.8)<br>p < 0.00 | $\Delta\beta = 14.5$<br>p < 0.00 | **0.00** |
| Motor Function/Dexterity | Pegboard NonDom Score | DWMH | -11.70 | $\beta = -3.73$ (95% CI: -7.85 to 0.394)<br>p 0.08 | $\beta = 5.82$ (95% CI: 3.45 to 8.18)<br>p < 0.00 | $\Delta\beta = 9.54$<br>p < 0.00 | **0.00** |
| Motor Function/Dexterity | Pegboard NonDom Score | Total WMH | -8.80 | $\beta = -4.42$ (95% CI: -12.5 to 3.7)<br>p 0.29 | $\beta = 9.62$ (95% CI: 6.42 to 12.8)<br>p < 0.00 | $\Delta\beta = 14$<br>p 0.00 | **0.00** |

Two-segment linear spline models showing differential WMH effects below and above test-specific thresholds. Thresholds were calculated from quadratic model inflection points using: $\text{knot} = -\beta_{\text{linear}} / (2 \times \beta_{\text{quadratic}})$ . 'Slope ≤ Threshold' represents $\beta$ per 1 log-unit WMH increase below the threshold; 'Slope $>$ Threshold' represents $\beta$ above the threshold. $\Delta\beta$ tests whether slopes significantly differ. Models adjusted for age, sex, race, education, systolic BP, and family clustering. q-values represent false discovery rate correction applied within each cognitive domain. N=339 for all tests except non-dominant hand pegboard tests (N=338). Significant slope differences suggest threshold effects where WMH impact accelerates above specific burden levels.

For PVWMH, the thresholds occurred at relatively low WMH burdens (log-transformed values ranging from −9.42 to −9.29). Below these thresholds, the relationship between PVWMH and pegboard performance was essentially flat or slightly protective (β coefficients ranged from −5.48 to −5.46, all p>0.05), suggesting minimal impact of low PVWMH burden on motor function. However, above the threshold, performance deteriorated dramatically, with slope coefficients indicating substantial increases in completion time (dominant hand time: β=9.62, p<0.001; nondominant hand time: β=9.58, p<0.001) and decreases in performance scores (dominant hand score: β=9.66, p<0.001; nondominant hand score: β=9.60, p<0.001). The differences in slopes between the two segments (Δβ) ranged from 14.5 to 15.1 (all p<0.001), confirming the presence of strong threshold effects.

DWMH demonstrated similar threshold patterns for motor function, though with thresholds at more negative log-transformed values (log-transformed values: −11.8 to −11.7) and somewhat smaller effect sizes. Below threshold, DWMH showed minimal associations with pegboard performance (all p>0.05), but above threshold, significant performance deterioration was evident (Δβ ranging from 9.54 to 10.9, all p<0.001). Total WMH volume showed comparable patterns, with threshold values intermediate between PVWMH and DWMH, and highly significant slope differences for all pegboard measures (Δβ ranging from 14.0 to 15.0, all p<0.001).

Processing speed, as measured by the digit symbol substitution test, demonstrated significant threshold effects for all three WMH subtypes. For correct responses, PVWMH showed a threshold at −11.3, below which higher WMH burden was paradoxically associated with better performance (β=6.68, p=0.001), but above which performance declined substantially (β=−4.48, p<0.001, Δβ=−11.2, p<0.001). DWMH demonstrated a similar bidirectional pattern with a threshold at −14.1 (Δβ=−9.77, p<0.001). These findings suggest that low WMH burden has minimal impact on processing speed, but cognitive efficiency deteriorates sharply once WMH exceeds critical levels.

Global cognitive function assessed by MMSE demonstrated a significant threshold effect for DWMH (threshold=−10.1, Δβ=−0.12, p=0.03), with minimal association below threshold (β=0.07, p=0.09) but the slope difference was significant (Δβ=−0.12, p=0.03). PVWMH and total WMH showed similar patterns, though the slope difference for PVWMH did not reach statistical significance (p=0.18), while total WMH showed a significant effect (p=0.03). These findings suggest that threshold effects on global cognition are present but less pronounced than those observed for motor and processing speed domains. Memory measures demonstrated variable threshold effects depending on WMH location. For short-term word recall, both PVWMH and total WMH showed threshold effects (PVWMH: Δβ=−0.27, p=0.10; total WMH: Δβ=−0.29, p=0.15), though not reaching statistical significance. DWMH demonstrated a nonsignificant threshold effect (threshold=−14.3, Δβ=−0.52, p=0.26). Long-term word recall showed similar patterns, with PVWMH demonstrating marginally significant threshold effects (Δβ=−0.59, p=0.05) and DWMH showing larger but non-significant effects. Executive function measures, including word fluency and digit span, showed selective threshold effects. Word fluency demonstrated a significant threshold effect for DWMH (threshold=−10.4, Δβ=−0.91, p=0.02), but not for PVWMH (p=0.49) or total WMH (p=0.29). Digit span forward showed a significant threshold effect for DWMH (threshold=−14.0, Δβ=1.18, p=0.001), while digit span backward demonstrated a non-significant threshold effect for PVWMH (threshold=−4.97, Δβ=−1.09, p=0.09).

Cross-sectional associations between WMH and cognitive performance at the 13-year follow-up were conducted as secondary, supplementary analyses to provide convergent support for the longitudinal threshold findings using the MOCA test only available at that time point. (see Supplemental Table 1)

## Discussion

This 13-year longitudinal study of 339 middle-aged adults from the GeneSTAR cohort employed two-segment linear spline mixed regression models to identify normalized lesion volume threshold effects in the relationships between changes in WMH burden and cognitive decline longitudinally. Our principal finding is that changing WMH does not have a linear effect on changing cognition. Cognitive decline accelerates markedly after exceeding specific inflection points in changing volume that varied by cognitive test. These thresholds estimated within each test-specific spline model represent the inflection points where the slope of cognitive decline changed most substantially. The magnitude of acceleration beyond these thresholds varied considerably by WMH anatomical location (periventricular versus deep) and cognitive test, with most pronounced accelerations observed for Grooved Pegboard (Δβ = 14.23-15.17, all p<0.001) and Digit Symbol Substitution Test (Δβ = −11.38, p<0.001). PVWMH showed stronger associations with cognitive decline than DWMH with pronounced effects on motor function. PVWMH and DWMH differ in overall volume distributions and absolute threshold values differ between them. Our normalization (WMH/ICV log-transformed) reduces scaling differences, and interpretation focuses on the strength of the associations rather than the threshold magnitude.

The use of two-segment linear spline models to characterize threshold effects is a methodological advance over traditional linear approaches that assume constant WMH effects across all burden levels. This strategy examining changes before and after specific transitions enabled us to identify the specific WMH burden levels at which cognitive decline accelerates. Below these thresholds, WMH burden showed minimal or even protective associations with cognitive performance. Above the thresholds performance deteriorated dramatically, confirming the existence of tipping points for declines in cognitive function.^36^

Our findings reveal differences in threshold effects by WMH anatomical location. PVWMH and DWMH demonstrated different thresholds depending on cognitive domains. PVWMH has larger threshold effects (Δβ: 14-15) for motor and processing speed. Periventricular regions include the corona radiata and internal capsule, which subserve motor and processing speed functions.^16^

The magnitude of threshold effects varied by cognitive test in different WMH locations, revealing domain-specific vulnerabilities. The largest threshold effects were observed for GPBT and DSS, both of which assess processing speed and psychomotor function. In contrast, MMSE demonstrated more modest threshold effects (DWMH Δβ = −0.147, p=0.013), while word recall measures showed variable patterns with larger effect sizes for DWMH (word recall short Δβ = −0.856, p=0.294) than PVWMH.^16^

Prior longitudinal studies show that individuals with a greater WMH burden at baseline exhibit faster cognitive decline over time. Our findings similarly demonstrate that baseline WMH burden predicts subsequent cognitive decline as in Puzo et al.^37^ Our study did not estimate annualized rates of decline, we demonstrate that changing WMH volume is associated with changing cognition in a nonlinear, threshold-dependent patterns.^37^ Baseline WMH volume did not independently predict conversion to MCI. WMH volume predicted change for each neuropsychological test score investigated independent of age. Examination of continuous neuropsychological test scores is perhaps optimal for the detection of brain behavior relationships, particularly in the absence or early stages of disease. This interaction captures subtle, continuous cognitive changes that precede categorical diagnostic shifts. Similarly, Carmichael et al.^38^ showed that baseline WMH independently predicted steeper 1-year declines in global cognition even after controlling for Alzheimer’s disease biomarkers.^38^

Our study advances this evidence by demonstrating that the predictive value of baseline WMH is not constant across all burden levels but rather concentrated above specific test-dependent thresholds. This has important implications for risk stratification: individuals with WMH burden just below threshold may experience minimal cognitive decline over extended follow-up periods, whereas those just above threshold face accelerated deterioration.

### Clinical Implications

Identification of test-specific and location-specific thresholds has important clinical implications. It challenges the common practice of treating WMH as a continuous risk factor with constant effects across all burden levels. Clinical risk calculators and prognostic models should incorporate threshold effects to avoid underestimating risk in individuals whose WMH burden has crossed critical thresholds.^39^ Secondly, the demonstration that PVWMH has larger threshold effects (Δβ: 14-15) on motor and processing speed functions than DWMH suggests that periventricular lesions may warrant closer attention in risk stratification and follow-up, particularly when PVWMH burden approaches or exceeds these thresholds, although current guidelines do not yet differentiate management by WMH location.^39^

Consistent with previous findings demonstrating WMH effects on manual dexterity in middle-aged adults,^11^ the dramatic acceleration of decline in Grooved Pegboard performance above threshold (slope increases of 14-15 seconds per log-unit WMH) has functional implications for activities of daily living requiring manual dexterity,^40^ such as dressing, cooking, and using mobile devices.^41^

### Strengths and Limitations

Major strengths of this study include the detailed phenotyping of vascular risk factors, extended 13-year follow-up, comprehensive neuropsychological battery, novel application of two-segment linear spline regression to detect threshold effects. The use of data-driven threshold identification from quadratic model inflection points avoided arbitrary cut point selection, while separate analyses for PVWMH, DWMH, and total WMH enabled anatomically specific inferences. All models were adjusted for major confounders, including age, sex, race, education, systolic blood pressure, and family clustering. Advanced imaging methods including deep learning-based WMH segmentation (UNISELF),^30^ longitudinal registration (Voxel Morph),^33^ and temporal lesion tracking ensured accurate quantification of progression.

Compared with prior WMH–cognition studies, our work provides a more rigorous and comprehensive evaluation of threshold effects. Earlier papers typically relied on either simple correlations, categorical severity scales, or single modeling strategies, which limit their ability to detect meaningful non-linearities.^17,42,43^ In contrast, our study uses normalized, log-transformed WMH volumes, evaluates periventricular, deep, and total WMH separately, and identifies thresholds using a fully data-driven two-step approach: quadratic-derived inflection points and independent grid-search validation using AIC/BIC. Additionally, our models adjust for demographics and vascular risk factors with family clustering and leverage a larger cognitive battery allowing us to map threshold behavior across multiple domains with greater precision.

Our study has some limitations, because the GeneSTAR cohort consists only of individuals with a family history of early coronary artery disease, the findings may not fully apply to people without this risk factor. At the same time, this is also a strength, it allows us to closely examine mechanisms of vascular cognitive impairment in a group at particularly high risk. As with any long-term study, there is a risk of attrition bias. Participants who passed away or developed severe cognitive problems over the 13 years are not included in later assessments causing underestimates of the true strength of these relationships. This study did not include biomarker data from blood or cerebrospinal fluid, which could have helped clarify underlying mechanisms and detect coexisting Alzheimer’s pathology or other dementing disorders.^44^ The 13-year interval between baseline and follow-up imaging required scanner replacement, with baseline acquisitions performed on a Philips 3T Achieva system and follow-up on an Ingenia system. This hardware evolution introduced potential technical variability despite harmonization procedures including N4 bias field correction^25^ and HACA3 contrast standardization.^29,30^ Scanner improvements generally enhance WMH detection sensitivity, potentially contributing to apparent progression in some participants. Consistency and magnitude of associations across multiple cognitive tests and analytical approaches suggest that observed effects reflect real biological phenomena.

### Future research directions

These results point toward several promising possibilities for future research. Including prospective validation studies targeted toward test-specific WMH thresholds in midlife. The observation that WMH progression accelerates approximately ten years before mild cognitive impairment onset suggests a critical window for intervention.^45^ Acting during midlife may offer the greatest benefit.^46^ Studying WMH regression mechanisms could identify modifiable factors helping tissue recovery. While WMH warrant intensive risk factor management when found incidentally,^47^ health economic analyses are needed to determine if systematic screening in at-risk populations is cost-effective.^48,49^

## Conclusions

This 13-year longitudinal study of middle-aged adults with elevated cardiovascular risk employed two-segment linear spline regression to demonstrate that changes in WMH volume effect changes in cognition over 13 years and are not linear over time and exhibit cognitive domain specific and location-specific thresholds beyond which cognitive decline accelerates markedly. The strongest threshold effects were observed for Grooved Pegboard (Δβ = 14.23-15.17, p<0.001) and Digit Symbol Substitution Test (Δβ = −11.38, p<0.001), with the greatest effect for PVWMH volume changes. These findings challenge the traditional view of WMH as a continuous risk factor with linear effects and highlight the importance of considering both anatomical location and magnitude of disease burden when assessing changing clinical risk. Because thresholds are associated with transitions from stable to accelerating cognitive decline, these results reveal the possibility of a targeted threshold to aid prevention strategies initiated in midlife. Further studies are needed to determine whether early detection and threshold-guided intensive interventions can alter trajectories of changing cognition in at-risk populations.

## Data Availability

Data are available from the GeneSTAR steering committee upon reasonable request and subject to data-use agreements.

## Abbreviations

cSVD: Cerebral small vessel disease
DSS: Digit Symbol Substitution
DWMH: Deep white matter hyperintensities
FDR: False Discovery Rate
FLAIR: Fluid-attenuated inversion recovery
GeneSTAR: Genetic Study of Atherosclerosis Risk
GPBT: Grooved Pegboard Test
HACA3: Harmonization with Attention-based Contrast, Anatomy, and Artifact Awareness
MMSE: Mini-Mental State Examination
MNI: Montreal Neurological Institute
MoCA: Montreal Cognitive Assessment
MPRAGE: Magnetization-prepared rapid gradient-echo
MRI: Magnetic resonance imaging
PVWMH: Periventricular white matter hyperintensities
SLANT: Spatially Localized Atlas Network Tiles
SMORE: Self-supervised Anti-aliasing and Super-resolution Algorithm)
TFE: Turbo field echo
UNISELF: Universal Self-supervised Learning Framework
WMH: White matter hyperintensities

## Author Contributions

P.A.N. Supervised the overall study. Designed and executed the study and aided with analysis and drafting of the article. S.S. Executed image processing, conceived, designed and performed statistical analyses and drafted the manuscript. L.K. supervised MRI acquisition protocols and imaging methodology and contributed to study design and neuroimaging interpretation. P.v.Z. supervised MRI acquisition protocols and imaging methodology. N.J.A. and D.V. provided statistical guidance and methodological consultation. M.M. recruited participants, executed the study and performed data entry. S.S., S.W., and J.Z. conducted imaging analysis and lesion segmentation. K.A.W. designed and validated the cognitive test battery. H.G.S. contributed to image processing methodology. R.L. provided clinical interpretation of imaging findings. J.L.P. supervised imaging analysis and algorithm development. L.C.B. and L.R.Y. designed and supervised the GeneSTAR cohort study and provided access to participant data. All authors critically reviewed and edited the manuscript and approved the final version.

## Funding

This study is funded by National Institutes of Health grants: NINDS/NIA award RF1NS128135. This research was supported, in part, by the Intramural Research Program of the National Institutes of Health (NIH). The contributions of the NIH author(s) are considered Works of the United States Government. The findings and conclusions presented in this paper are those of the author(s) and do not necessarily reflect the views of the NIH or the U.S. Department of Health and Human Services.

## Competing interests

KAW is an Associate Editor for Alzheimer’s & Dementia: The Journal of the Alzheimer’s Association, Alzheimer’s & Dementia: Translational Research and Clinical Interventions (TRCI), and on the Editorial Board of Annals of Clinical and Translational Neurology. KAW is on the Board of Directors of the National Academy of Neuropsychology. KAW has given unpaid presentations and seminars on behalf of SomaLogic.

## Supplemental Material

Table S1

Figure S1

## Notes

### Clinical Trial

Not applicable, this is an observational cohort study and not a clinical trial.

### Author Declarations

Approved by Johns Hopkins University IRB, protocol NA_00002856.

